# Selection, optimization, and validation of ten chronic disease polygenic risk scores for clinical implementation in diverse populations

**DOI:** 10.1101/2023.05.25.23290535

**Authors:** Niall J Lennon, Leah C Kottyan, Christopher Kachulis, Noura Abul-Husn, Josh Arias, Gillian Belbin, Jennifer E Below, Sonja Berndt, Wendy Chung, James J. Cimino, Ellen Wright Clayton, John J. Connolly, David Crosslin, Ozan Dikilitas, Digna R. Velez Edwards, QiPing Feng, Marissa Fisher, Robert Freimuth, Tian Ge, The GIANT Consortium, The All of Us Research Program, Joseph T. Glessner, Adam Gordon, Candace Guiducci, Hakon Hakonarson, Maegan Harden, Margaret Harr, Joel Hirschhorn, Clive Hoggart, Li Hsu, Ryan Irvin, Gail P. Jarvik, Elizabeth W. Karlson, Atlas Khan, Amit Khera, Krzysztof Kiryluk, Iftikhar Kullo, Katie Larkin, Nita Limdi, Jodell E. Linder, Ruth Loos, Yuan Luo, Edyta Malolepsza, Teri Manolio, Lisa J. Martin, Li McCarthy, James B Meigs, Tesfaye B. Mersha, Jonathan Mosley, Bahram Namjou, Nihal Pai, Lorenzo L. Pesce, Ulrike Peters, Josh Peterson, Cynthia A. Prows, Megan J. Puckelwartz, Heidi Rehm, Dan Roden, Elisabeth A. Rosenthal, Robb Rowley, Konrad Teodor Sawicki, Dan Schaid, Tara Schmidlen, Roelof Smit, Johanna Smith, Jordan W. Smoller, Minta Thomas, Hemant Tiwari, Diana Toledo, Nataraja Sarma Vaitinadin, David Veenstra, Theresa Walunas, Zhe Wang, Wei-Qi Wei, Chunhua Weng, Georgia Wiesner, Yin Xianyong, Eimear Kenny

## Abstract

Polygenic risk scores (PRS) have improved in predictive performance supporting their use in clinical practice. Reduced predictive performance of PRS in diverse populations can exacerbate existing health disparities. The NHGRI-funded eMERGE Network is returning a PRS-based genome-informed risk assessment to 25,000 diverse adults and children. We assessed PRS performance, medical actionability, and potential clinical utility for 23 conditions. Standardized metrics were considered in the selection process with additional consideration given to strength of evidence in African and Hispanic populations. Ten conditions were selected with a range of high-risk thresholds: atrial fibrillation, breast cancer, chronic kidney disease, coronary heart disease, hypercholesterolemia, prostate cancer, asthma, type 1 diabetes, obesity, and type 2 diabetes. We developed a pipeline for clinical PRS implementation, used genetic ancestry to calibrate PRS mean and variance, created a framework for regulatory compliance, and developed a PRS clinical report. eMERGE’s experience informs the infrastructure needed to implement PRS-based implementation in diverse clinical settings.

## Introduction

Polygenic risk scores (PRS) are being calculated and disseminated at a prodigious rate[1], but their development and application to clinical care, particularly among ancestrally diverse individuals, present substantial challenges[2–4]. Incorporation of genomic risk information has the potential to improve risk estimation and management [3,5], particularly at younger ages [6]. Clinical use of PRS may ultimately prevent disease or enable its detection at earlier, more treatable stages [6–8]. Improved estimation of risk may also enable targeting of preventive or therapeutic interventions to those most likely to benefit from them while avoiding unnecessary testing or over-treatment [9,10].

PRS for individual conditions are typically generated from summary statistics derived from genome-wide association studies (GWAS), which are themselves derived from populations that are heavily over-represented by individuals of European ancestry [11]. Such scores have been shown to have limited prediction accuracy with increasing genetic distance from European populations [11,12]. PRS can be improved if developed and validated using multi-ancestry cohorts [13]. Clinical and environmental data combined with genomically-derived risk measurements can improve risk prediction [14]. Approaches for combining genomic and non-genomic information, optimizing models for genomically diverse populations and across age groups, and conveying this information to clinicians and patients have yet to be developed and applied in clinical care.

The Electronic Medical Records and Genomics (eMERGE) Network is a multicenter consortium established in 2007 to conduct genomic research in biobanks with electronic medical records [15,16]. In 2020, eMERGE embarked on a study of genomic risk assessment and management in 5,000 children and 20,000 adults of diverse ancestry, beginning with efforts to identify and validate published PRS across multiple race-ethnic groups (and inferred genetic ancestries) in 10 common diseases with complex genetic etiologies. This paper describes identification, selection, and optimization of these PRS; calibration of ancestry for PRS estimation using a novel method developed for eMERGE; and development and launch of clinical reporting tools.

## Results

### PRS Auditing and Evaluation

To select the PRS for clinical implementation, the network conducted a multi-stage process to evaluate proposed scores (Figure 1). An initial set of 23 conditions was selected based on considerations including relevance to population health (condition prevalence and heritability), strength of evidence for PRS performance, clinical expertise in the eMERGE Network, and data availability that would facilitate validation of the PRS in diverse populations. Network sites completed comprehensive literature reviews on 23 proposed conditions and the corresponding PRS. A summary of the features of the PRS for each of the final conditions chosen is shown in Supplemental Table 1. The collated information included *analytic viability* - a description of covariates, the age, and ancestry effects of the original PRS model; *feasibility* - access to sufficiently diverse validation data sets (race/ethnicity and age) as well as condition prevalence and relevance to preventative care; *potential clinical actionability* - existing screening or treatment strategies, and magnitude (odds ratio) of risk in the high risk group; and *translatability* - expected public health impact across diverse populations. Candidate PRS were restricted to those that were either previously validated and published (journal or pre-print) or for which there was sufficient access to information to develop and/or optimize new PRS, which could then be validated.

**Figure 1.**
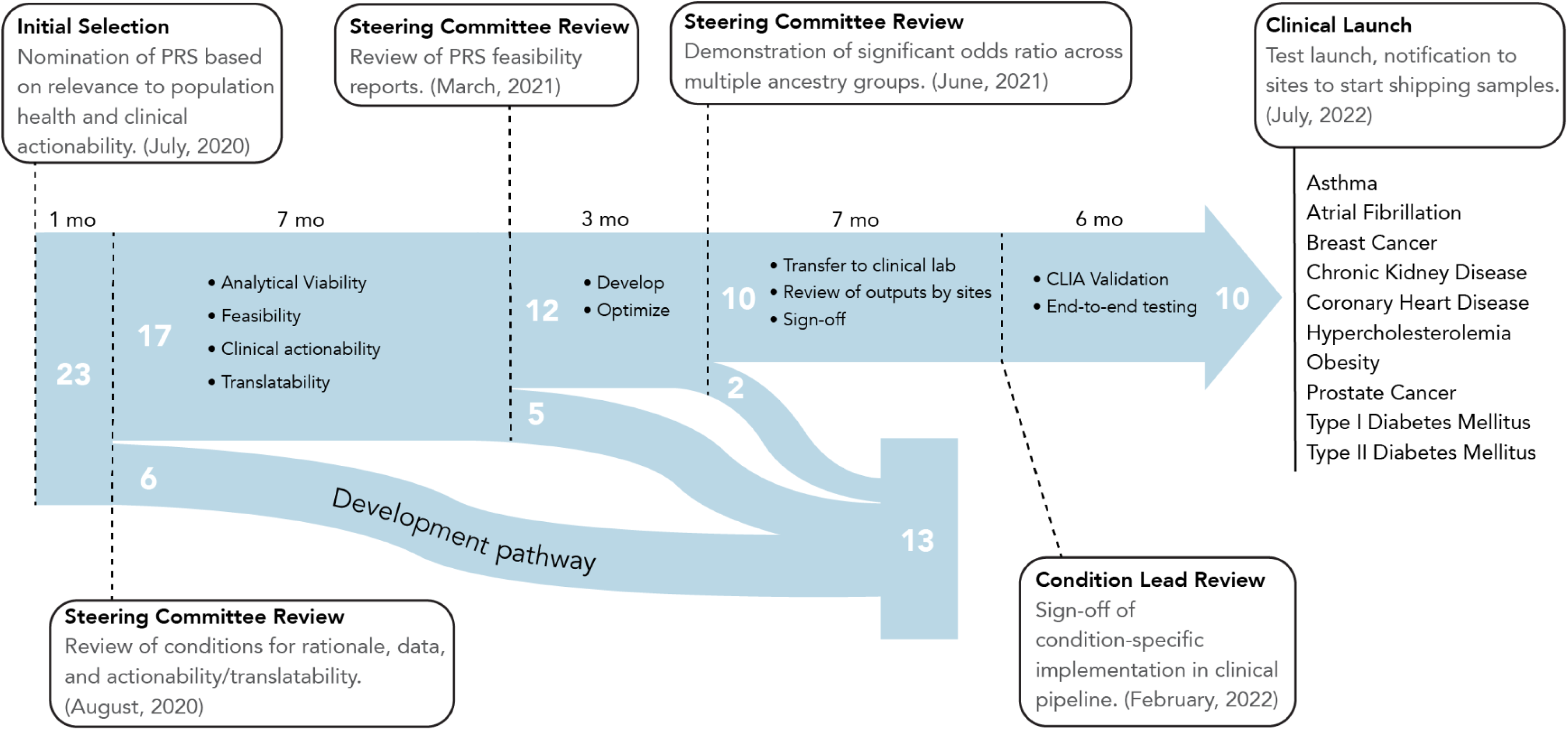
Timeline and process for selection, evaluation, optimization, transfer, validation, and implementation of the clinical PRS test
pipeline. Dotted lines represent pivotal moments in the progression of the project.

**Figure 2.**
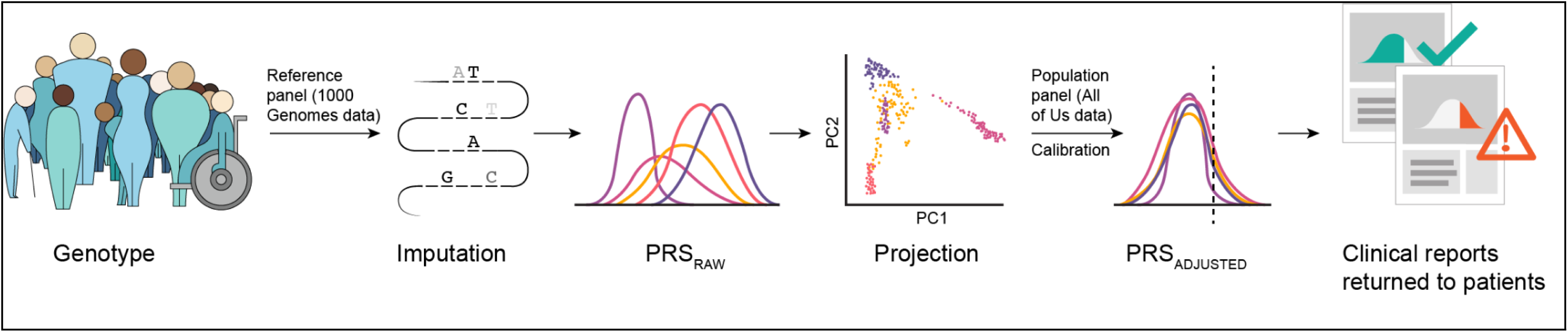
Overview of the eMERGE PRS process. Participant DNA is genotyped using the Illumina Global Diversity Array which assesses 1.8M sites. Genotyping data is phased and imputed with a reference panel derived from the 1000 Genomes Project. Raw PRS scores are calculated for each condition. For each condition an ancestry calibration model is applied based on model parameters derived from the All of Us Research Program. Participants whose adjusted scores cross the pre-defined threshold for high PRS are identified and a pdf report is generated. The report is electronically signed after data review by a clinical laboratory director and delivered to the study portal for return to the clinical sites.

In auditing and evaluating evidence of PRS performance, the eMERGE Steering Committee (SC) considered PRS for conditions that could be implemented in pediatric and/or adult populations, and for diseases with a range of age of onset (0 to >65 years of age). We considered published SNP-based heritability estimates available for 10 of the 23 conditions, ranging from 3% to 58%. The majority of PRS under consideration aimed to identify individuals at high risk for disease; however, PRS to predict disease severity and drug response were also considered. Two of the conditions, breast cancer and prostate cancer, were only considered for implementation in individuals whose biological sex was female or male, respectively. As the eMERGE network plans to enroll >50% participants from underrepresented groups (including racial and ethnic minority groups; people with lower socioeconomic status (SES); underserved rural communities; sexual and gender minority (SGM) groups) [17] emphasis was placed on the PRS that were already available for, or could be developed and validated in, diverse population groups.

To define population groups, study-level population descriptors were first extracted from published literature, pre-prints or information shared directly by collaborators on data used to develop and/or optimize and/or validate PRS. Methods for using population labels across studies ranged from self-reporting, extraction from health system data, and/or analysis of genetic ancestry. We designated four population groups; European ancestry (EA) (i.e. study population descriptors included European, European-American, or other European descent diaspora groups), African (African, African-American (AA), or other African descent diaspora groups), Hispanic (HL) (i.e. Hispanic, Latina/o/x, or those who have origins in countries in the Caribbean and Latin America), and Asian (Asn) (i.e. South Asian, East Asian, South-East Asian, Asian-American or other diaspora Asian groups).

Of the 23 conditions initially selected, six were excluded at the outset (August 2020) due to lack of diversity in the PRS training or optimization data, lack of access to diverse datasets for validation, or lack of available clinical expertise in the network (Figure 1). A further four conditions were dropped in March 2021 due to insufficient confidence in PRS performance or lack of validation datasets. Conditions not prioritized for implementation continued on a ‘developmental’ pathway for further refinement. Each of the 12 conditions that were selected to move forward from the March 2021 review were assigned a ‘lead’ and ‘co-lead’ site which worked together to develop, validate, and transfer the score to the clinical laboratory for instantiation and CLIA validation. Assignment of leads was based on site preference, expertise, and distribution of workload.

### Selection, optimization, and validation

A systematic framework was developed to evaluate the performance for the remaining 12 PRS, in accordance with best practices outlined in Wand *et al* [18]. An in depth evaluation matrix of the 12 chosen conditions can be found in Supplemental Table 2. Clinical use of Eurocentric PRS in diverse patient samples risks exacerbating existing health disparities[11][19,20]. The Network carefully considered a variety of strategies to optimize PRS generalizability and portability. The Network prioritized validation across four ancestries with an emphasis on African and Hispanic ancestry due to their underrepresentation in genetic research and projected representation within the study cohort. We determined that a PRS was validated if the genomic predictor was significantly discriminative and the odds ratios were statistically significant in a minimum of two and up to four ancestral populations: African/African-American, Asian, European Ancestry; and Hispanic/Latino. The PRS Working Group members conducted an extensive scoping exercise to identify suitable datasets of multiple ancestries for disease-specific PRS validation. These included datasets from early phases of eMERGE (2007-2019) as well as external datasets such as the UK Biobank and Million Veteran Program (MVP). A standardized set of questions were addressed by the disease leads that included the source of discovery and validation datasets, the availability of multi-ancestry validation datasets, availability of cross-ancestry PRS, proposed percentile thresholds for identifying high risk status, model discrimination (AUC), and effect sizes (odds ratios) associated with high risk vs. not-high risk status (Supplemental Table 2). For 7 out of the 12 candidate scores, no further optimization of the original model was performed. For 5 scores, an additional optimization effort was undertaken to further refine the score performance in multiple ancestries. Details of the optimization can be found in Supplemental Table 3. A specific score optimization was applied for CKD. This optimization consisted of adding the effect of APOL1 risk genotypes to a polygenic component, which has been found to improve risk predictions in African ancestry cohorts[21].

For final selection, the steering committee considered the score performance summaries (presented by condition leads) in addition to the actionable and measurable recommendations relevant for return, for each condition, in the prospective cohort. Two conditions (colorectal cancer (CRC) and abdominal aortic aneurysm (AAA)) were moved to the developmental pathway (Figure 1). While the PRS for CRC was not included in the prospective cohort, as several Mendelian genes for Lynch syndrome were included in the custom panel generated by Invitae, the network decided to include CRC in the list of conditions returned with the overall risk report (GIRA) and only report on monogenic and family history risk.

### Population-based z-score calibration

In this study, the focus is on integration and implementation of validated PRS in clinical practice rather than novel PRS development. Ultimately, the Network opted to balance generalizability and feasibility by validating and returning cross-ancestry PRS. However, even with cross-ancestry scores, differences remain in the distribution of z-scores across genetic ancestries that can result in inconsistent categorization of individuals into ‘high’ or ‘not high’ polygenic risk categories for a given condition [22]. To that end, the Network chose to develop methods to determine each participant’s ancestry and calibrate the distribution of resulting z-scores through a population-based calibration model[22][23] (see below). An alternative would have been to apply existing PRS in available samples of different ancestries and derive ancestry-specific effect estimates. However, returning ancestry-specific risk estimates is challenging in real world implementations as it would require self-reporting of ancestry by patients (who may not be able to provide this with accuracy) and developing multiple ancestry-specific reports for each health condition. In addition, such PRS would be problematic to return to patients of mixed ancestry.

Polygenic risk scores often have different mean and standard deviation for individuals from different genetic ancestries. While some of these differences could be due to true biological differences in risk, they also result from allele frequency and linkage disequilibrium (LD) structure differences between populations [24]. This problem is more acute when a PRS is calculated for an individual whose ancestry does not match the ancestries used to develop the PRS. A clinically implemented PRS test to return disease risk estimates, therefore, must be adjusted to account for these differences due to ancestral background. A calibration method based on principal component analysis (PCA) which was initially described by Khera *et al*. [22] was modified to model both the variance and means of scores as ancestry dependent, as compared to the previous method, which modeled only the means as dependent on ancestry. This modification was found to be necessary because some conditions were found to exhibit highly ancestry-dependent variance (see for example, Figure 3 in Online Methods), which would have led to many more or fewer participants of certain ancestries receiving a ‘high PRS risk’ determination than was intended. The model was fit to a portion of the All of Us Research Program (https://www.researchallofus.org/) cohort genotyping data, which allowed for continuous return of results to participants without needing to wait for the entire study dataset to be available. More details can be found in Online Methods.

**Figure 3.**
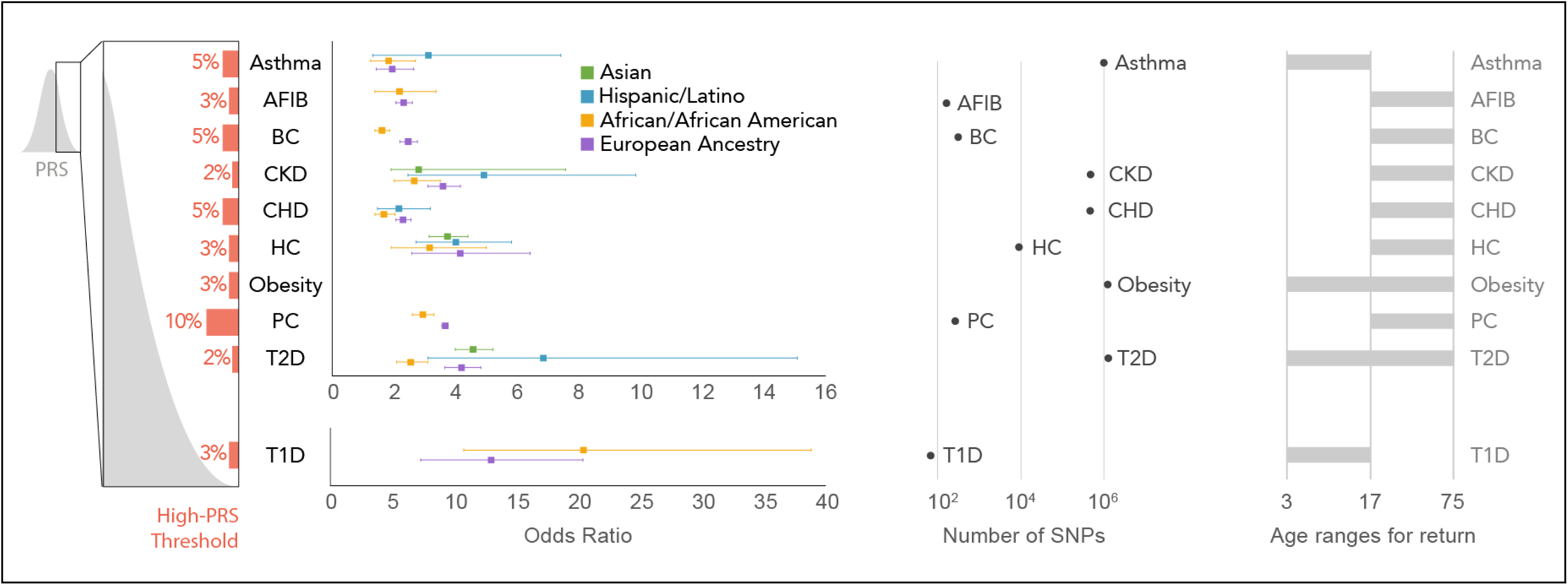
Summary of the ten conditions that were implemented. “High-PRS Threshold” represents the percentile that is deemed to be the cut-off for a specific condition above which a high PRS result is reported for that condition. The Odds Ratios are the OR of the implemented scores, 95% confidence interval shown in the whiskers (with the exception of Obesity for which the OR will be published by the GIANT consortium). “Number of SNPs” represents the range of numbers or sites included in each score. “Age ranges for return” indicates the participant ages at which a PRS is calculated for a given condition. AFIB= Atrial fibrillation; BC = Breast Cancer; CKD = Chronic Kidney Disease; CHD = Coronary Heart Disease; HC = Hypercholesterolemia; PC = Prostate Cancer; T2D = Type 2 Diabetes; T1D = Type 1 Diabetes.

### Transfer and Implementation

Once the final 10 conditions had been selected, condition-leads worked with computational scientists at the clinical laboratory (Clinical Research Sequencing Platform, LLC at the Broad Institute) to transfer the PRS models. Condition-specific models were run with outputs from the lab’s genotyping (Illumina Global Diversity Array), Phasing (Eagle2 [25] https://github.com/poruloh/Eagle), and imputation (Minimac4 [26] https://genome.sph.umich.edu/wiki/Minimac4) pipelines to assess genomic site representation (see Online Methods for more information on the architecture and components of the pipeline). Several rounds of iteration between the clinical laboratory and condition-leads followed in which any issues with the pipeline were resolved and the effect of genomic site missingness was assessed (Table 1). The final version of the implemented models was returned to the condition leads to recalculate effect sizes in the validation cohorts.

**Table 1.**
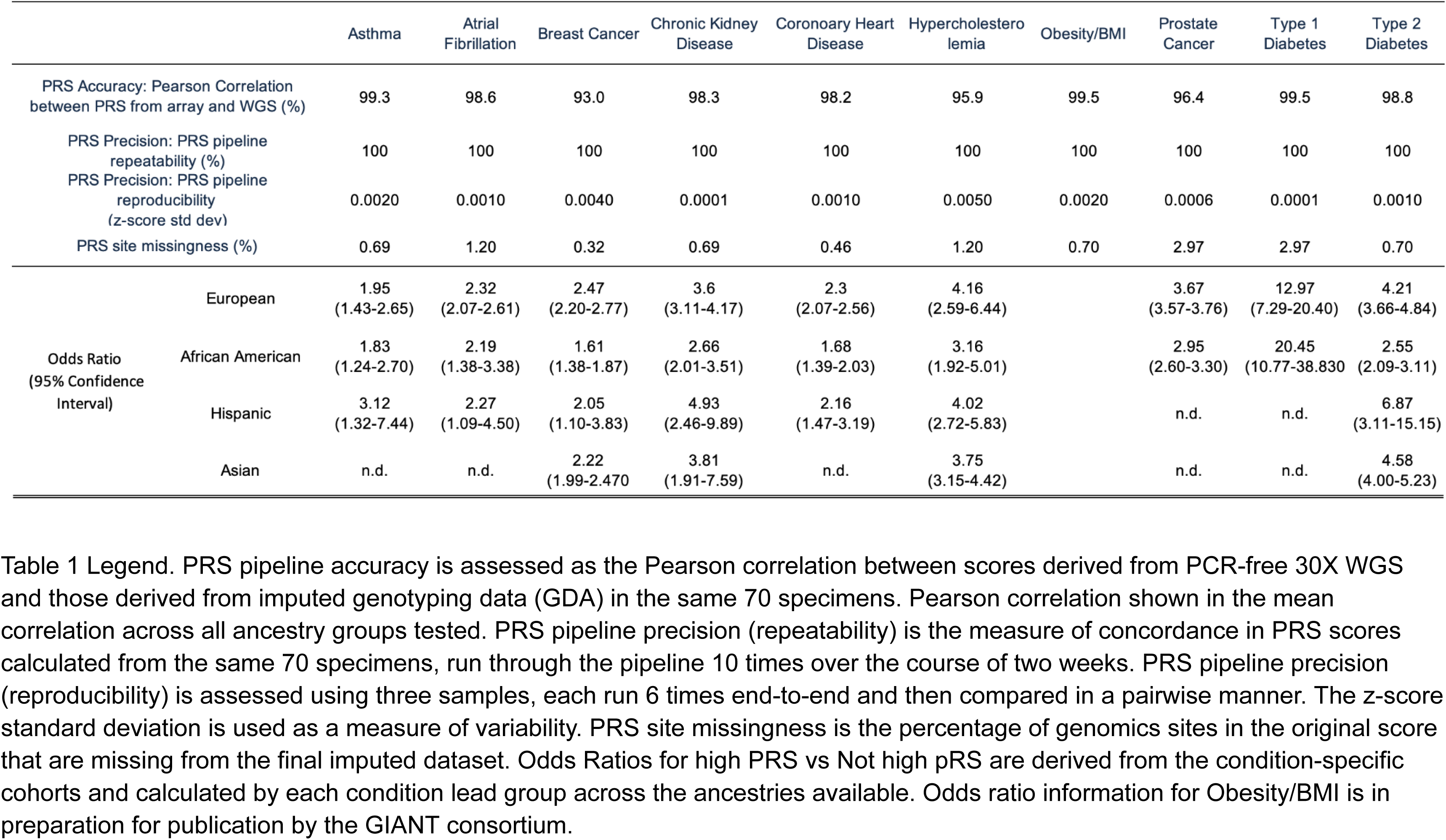
Performance measures from the PRS pipeline validation study at the clinical laboratory.

Finally, as part of the implementation of the PRS pipelines as a clinical test in a CLIA laboratory, a validation study was performed (See Online Methods for a detailed description, Table 1 summarizes some of the results). Briefly, this study leveraged 70 reference cell lines from diverse ancestry groups (Coriell) where 30X whole genome sequencing data was generated to form a variant truth set from which the technical accuracy and reproducibility of imputation and PRS calling was assessed. A second sample set of 20 matched donor blood and saliva specimens was procured to assess the performance of the pipeline with different input materials. A set of three samples, each with 6 replicates, was run end-to-end through the wet lab and analytical pipelines as an assessment of reproducibility. As a verification of the clinical validity of the scores, cohorts of cases for 8 of the 10 conditions were created using the eMERGE phase III imputed dataset (available on https://anvil.terra.bio/#workspaces/anvil-datastorage/AnVIL_eMERGE_GWAS/data (registration required)). PRS performance measures were calculated to confirm associations between scores and conditions. Due to limitations in the eMERGE phase III imputation (no chromosome X, different imputation pipeline) the ORs from this analysis were not included in the final reports, rather the ORs calculated in the condition-specific validation cohorts (using the final clinical lab pipeline) were used (Figure 3 and Table 1). A validation report was created for each condition. This report was reviewed and approved by the Laboratory Director in compliance with CLIA regulations for the development of a laboratory developed test. Personnel were trained on laboratory and analytical procedures, and standard operating procedures were implemented. Data review metrics were established, sample pass/fail criteria were defined, and order and report data transfer pipelines were built as described in Linder *et al.* [27]

### Creation of report and pipeline for report creation, review, sign-out, release

A software pipeline was built to facilitate data review and clinical report generation in both document (pdf) and structured data formats (sample report included in Supplementary Material). Logic was built into the PRS and reporting pipeline to account for differences in return based on age and sex at birth for certain conditions. For instance, the PRS for breast cancer is only calculated for participants who report sex at birth as female; similarly prostate cancer scores are only generated for participants who report sex at birth as male. Age-related restrictions were similarly coded into the pipeline to account for study policies on return. Data review by an appropriately qualified, trained individual is required for high complexity clinical testing. In the PRS clinical pipeline this review takes the form of a set of metrics that are exposed by the pipeline to the reviewer. These include a z-score range for each condition (passing samples will have a score -5 < z < +5), a PCA plot per batch against a reference sample set (visual representation of outlier samples), monitoring the z-score range for each control per condition (one control on each plate; NA12878), and flagging any samples with multiple ‘High Risk’ results for further review.

Each participant’s sample is also run on an orthogonal fingerprinting assay (Fluidigm biomark) that creates a genotype-based fingerprint for that DNA aliquot. Infinium genotyping data is compared to this fingerprint as a primary check of sample chain-of-custody fidelity and to preclude sample or plate swaps during lab processing.

Reviewed and approved data for a participant is processed into a clinical report. The text and format of this report were created during an iterative review process by consortium work groups. For this pragmatic clinical implementation study, two results are returned to participants: “High Risk” or “Not High Risk” based on the PRS [27]. In the clinical report a qualitative framework has been developed to indicate for which condition(s) a participant has been determined to have a high PRS (if any). Quantitative values (Z-scores) are not included for any condition in the main results panel. For breast cancer and CHD, the z-score is presented in another section of the report for inclusion in integrated score models for those conditions. For breast cancer specifically, the provided z-score is used with the BOADICEA[28] model to generate an integrated risk that is included in the genome-informed risk assessment (GIRA) as described in Linder *et al*.[27]

### Overview of first 2500 clinical samples processed

Between launch in July 2022 and May 2023, 2500 participants have been processed through the clinical PRS pipeline (representing ∼10% of the proposed cohort). Of the first 2500 participants processed, 64.5% (1612) indicated sex at birth as female, while 35.5% (886) indicated male. Median age at sample collection was 51 years (range 3 years to 75 years). Participants self-reported race/ancestry, with 32.8% (820) identifying as “White (e.g. English, European, French, German, Irish, Italian, Polish, etc)”; 32.8% (820) identified as “Black, African American, or African (e.g. African American, Ethiopian, Haitian, Jamaican, Nigerian, Somali, etc.)”; 25.4% (636) identified as “Hispanic, Latino, or Spanish (e.g. Colombian, Cuban, Dominican, Mexican or Mexican American, Puerto Rican, Salvadoran, etc.)”; 5% (124) identified as “Asian (e.g. Asian, Indian, Chinese, Filipino, Japanese, Korean, Vietnamese, etc.)”; 1.5% (38) identified as American Indian or Alaska Native (e.g. Aztec, Blackfeet Tribe, Mayan, Navajo Nation, Native Village of Barrow (Utqiagvik) Inupiat Traditional Government, Nome Eskimo Community, etc.); 0.9% (22) identified as Middle Eastern or North African (e.g. Algerian, Egyptian, Iranian, Lebanese, Moroccan, Syrian, etc.); 0.8% (21) selected “None of these fully describe [me_or_my_child]”; 0.7% (17) selected “Prefer not to answer”; 0.1% (2) participants had incomplete data. A summary of the performance of the first 2500 samples and resulting high PRS metrics are shown in Figure 4. In the first 2500 participants, we identified 515 participants (20.6%) with a high PRS risk for one of the 10 conditions, 64 participants (2.6%) had high PRS risk for two conditions, and two participants (0.08%) had a high risk for three conditions. The remaining 1919 participants had no high PRS found. High PRS participants spanned the spectrum of genetic ancestry when projected onto principal component space (Figure 4).

**Figure 4.**
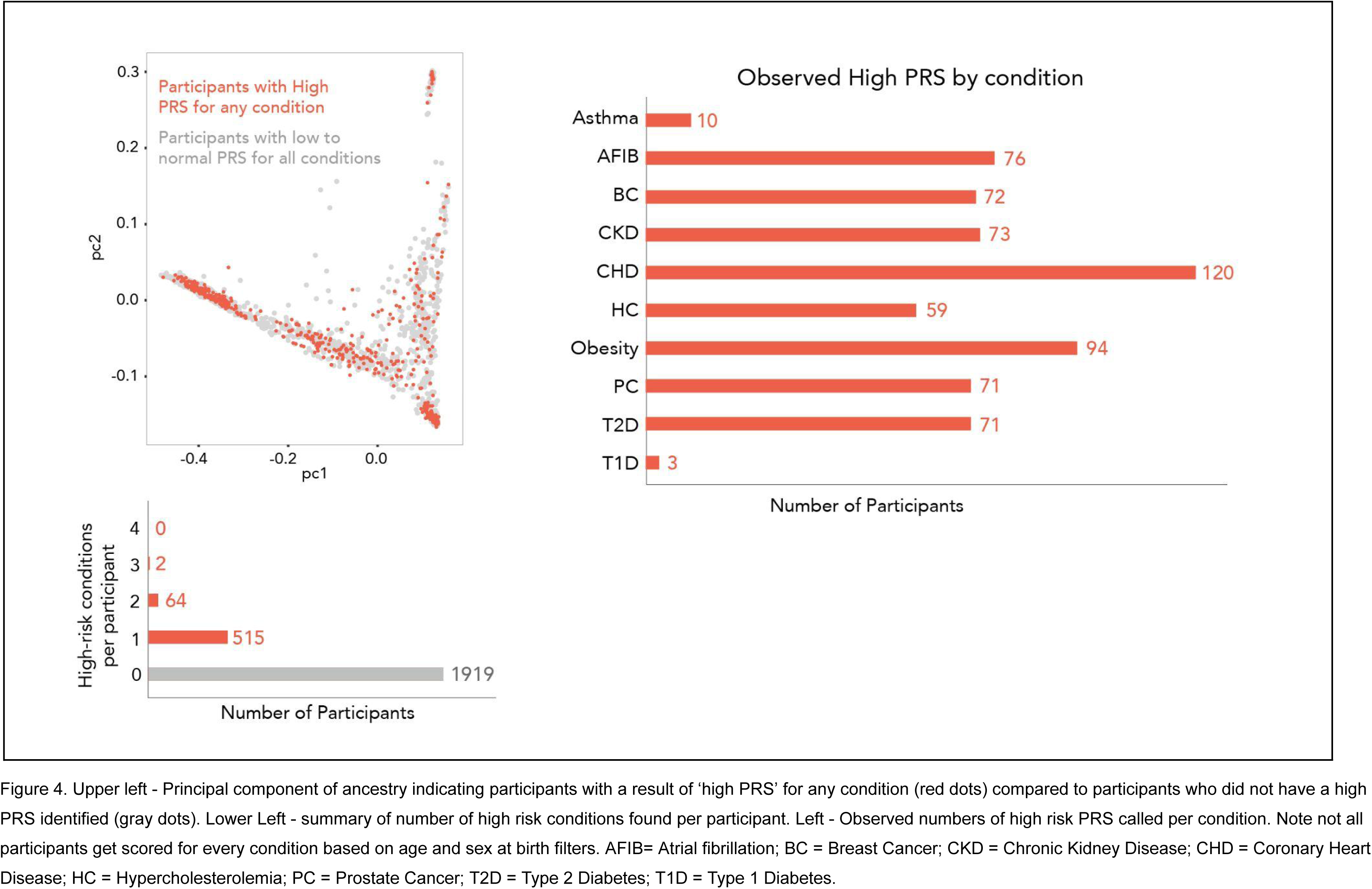
Summary of first 2500 clinical samples

## Discussion

While the predictive performance of PRS has improved significantly in recent years, challenges remain in ensuring that PRS are applicable and effective in diverse populations. In particular, the vast majority of GWAS have focused on individuals of European ancestry, and the predictive accuracy of PRS declines with increasing genetic distance from the discovery population[29][24][4]. This risks exacerbating existing health disparities, as clinical use of Eurocentric PRS in diverse patient samples may not accurately reflect disease risk in non-European populations. To address these challenges, the eMERGE Network has conducted a multi-stage process to evaluate and optimize PRS selection, development, and validation. The network has prioritized conditions with high prevalence and heritability, existing literature, clinical actionability, and the potential for health disparities, and has developed strategies to optimize PRS generalizability and portability across diverse populations. In particular, the network has emphasized performance across four major ancestry groups (African, Asian,

European, Hispanic, as reflected by self-identified race/ethnicity) and has developed a pipeline for clinical PRS implementation, a framework for regulatory compliance, and a PRS clinical report. The potential impact of PRS-based risk assessment in clinical practice is significant. By enabling targeted interventions and preventative measures, PRS-based risk assessment has the potential to reduce the burden of a range of conditions [27]. Moreover, the development of PRS-based risk assessment in diverse populations has the potential to reduce health disparities by ensuring that clinical use of PRS accurately reflects disease risk in diverse populations.

However, challenges remain in the successful implementation of PRS-based risk assessment in clinical practice. These include concerns about genetic determinism, the potential for stigmatization, and the need for robust regulatory frameworks to ensure that PRS-based risk assessment is deployed safely and effectively. Additionally, one of the biggest challenges is the implementation of effective disease prevention strategies after the return of the results. Return of the results won’t result in a benefit without effective disease prevention or early detection strategies. The eMERGE Network’s work provides a promising blueprint for addressing these challenges, but ongoing research and evaluation will be necessary to ensure that PRS-based risk assessment is implemented in a responsible and effective manner.

In conclusion, the eMERGE Network’s work in PRS development represents a significant step forward in the implementation of PRS-based risk assessment (in combination with other risk estimates from monogenic testing and family history) in clinical practice. By leveraging the power of genetics to predict disease risk and enable targeted interventions, genetically-informed risk assessment has the potential to revolutionize personalized medicine and usher in a new era of precision health. While challenges remain in ensuring that PRS are applicable and effective in diverse populations, the eMERGE Network’s work provides a promising foundation for the continued development and evaluation of PRS-based risk assessment in clinical practice.

## Data Availability

Data produced are available online for registered users at https://anvilproject.org/

## Acknowledgements

The authors would like to thank the past and future participants of the eMERGE network projects. The authors would like to thank Mary O’Reilly for help with figure creation. We thank our All of Us Research Program colleagues, Andrea Ramirez, Sokny Lim, Brandy Mapes, Ashley Green and Anji Musick for providing their support and input throughout the ancestry calibration demonstration project lifecycle. We also thank the All of Us Science Committee and All of Us Steering Committee for their efforts evaluating and finalizing the approved demonstration projects. The All of Us Research Program would not be possible without the partnership of contributions made by its participants. To learn more about the All of Us Research Program’s research data repository, please visit https://www.researchallofus.org/.

## Author Information

These Authors contributed equally Niall Lennon, Leah C.Kottyan

## Authors and Affiliations

**Broad Institute of MIT and Harvard**

Marissa Fisher, Candace Guiducci, Maegan Harden, Chris Kachulis, Amit Khera,Katie Larkin, Niall J. Lennon, Edyta Malolepsza, Li McCarthy, Nihal Pai, Heidi Rehm, Diana Toledo

**Children’s Hospital Boston**

Joel Hirschhorn

**Children’s Hospital of Philadelphia**

John J. Connolly, Joseph T. Glessner, Hakon Hakonarson, Margaret Harr

**Cincinnati Children’s Hospital Medical Center**

Leah C. Kottyan, Lisa J. Martin, Tesfaye B. Mersha, Bahram Namjou, Cynthia A. Prows

**Columbia University**

Wendy Chung, Atlas Khan, Krzysztof Kiryluk, Chunhua Weng

**Fred Hutchinson Cancer Center and University of Washington**

Li Hsu, Ulrike Peters

**Icahn School Mount Sinai**

Gillian Belbin, Clive Hoggart, Roelof Smit, Zhe Wang

**Invitae**

Tara Schmidlen

**Mass General Brigham**

Tian Ge, Elizabeth W. Karlson, Elizabeth W. Karlson, James B Meigs, Jordan W. Smoller

**Mayo Clinic**

Ozan Dikilitas, Robert Freimuth, Iftikhar Kullo, Dan Schaid, Johanna Smith

**National Institutes of Health**

Josh Arias, Sonja Berndt, Teri Manolio, Robb Rowley

**Northwestern University**

Adam Gordon, Yuan Luo, Lorenzo L. Pesce, Megan J. Puckelwartz, Konrad Teodor Sawicki, Theresa Walunas

**University of Alabama at Birmingham**

James J. Cimino, Ryan Irvin, Nita Limdi, Hemant Tiwari

**University of Copenhagen**

Ruth Loos

**University of Michigan**

Yin Xianyong

**University of Washington**

David Crosslin, Minta Thomas, David Veenstra

**University of Washington Medical Center**

Gail P. Jarvik, Elisabeth A. Rosenthal

**Vanderbilt University Medical Center**

Jennifer E Below, Ellen Wright Clayton, Digna R. Velez Edwards, QiPing Feng, Jodell E. Linder, Jonathan Mosley, Josh Peterson, Dan Roden, Nataraja Sarma Vaitinadin, Wei-Qi Wei, Georgia Wiesner

**The GIANT Consortium**

**The All of Us Research Program**

## Online Methods/Supplemental Information Clinical PRS Paper

### A. Analytical and Technical Validation Studies

#### Broad Imputation Pipeline Overview

An imputation pipeline that takes as an input a variant call format (VCF) file generated from a genotyping microarray and imputes the genotypes at additional sites across the genome was developed. The pipeline architecture and composition was based on the widely used University of Michigan Imputation Server which uses a software called *Eagle* (https://github.com/poruloh/Eagle) for phasing and *Minimac4* (https://genome.sph.umich.edu/wiki/Minimac4) for the imputation. The pipeline uses a curated version of the 1000 Genomes Project (1KG, www.internationalgenome.org) as the reference panel. Additional details on the imputation pipeline can be found at https://broadinstitute.github.io/warp/docs/Pipelines/Imputation_Pipeline/README.

#### Broad Curated 1KG Reference Panel

During the validation process we determined that some sites in the 1KG reference panel were incorrectly genotyped compared to the sites in matching whole-genome sequencing data. In order to increase accuracy of the imputation and PRS scoring, we curated the original panel by removing sites that were likely incorrectly genotyped based on comparing allele frequencies to those reported in gnomAD v2 (https://gnomad.broadinstitute.org/). Documentation of this curation can be found at: https://broadinstitute.github.io/warp/docs/Pipelines/Imputation_Pipeline/references_overview and a publicly available version of the panel at: gs://broad-gotc-test-storage/imputation/1000G_reference_panel/

Selection of a reference panel for imputation as an input to PRS is an important consideration. Some reference panels (e.g. TOPMed) have more samples than the default used in our pipeline (i.e. 1KG). This leads to more variants being imputed. The question is whether this would materially change the PRS calculated from samples imputed with the TOPMED panel. Access to this panel computationally is restricted (and local download prohibited) so it was deemed infeasible to implement in our clinical production environment. The performance of a non-eMERGE PRS (for CHD, Khera et al.) using the two different reference panels was determined for 20 GDA saliva specimens and for 42 AoU v1 specimens. The cohort was imputed both by the Broad imputation pipeline with curated 1KG as the reference panel as well as on the TOPMed Imputation Server with TOPMed as the reference panel. Imputed arrays were scored by the PRS pipeline.

The PRS percentiles computed with each method are highly concordant for both cohorts. The Pearson correlation coefficient is 0.996 for both cohorts, the p-value of the Welch two sample t-test is equal to 0.93 and 0.85 (indicating no statistical difference between the methods) for GDA and AoU v1 cohorts, respectively.

##### Performance verification of the Imputation Pipeline

Imputation accuracy was determined for 42 specimens that were processed through a genotyping microarray (AoU v1 array - the precursor to the commercial Global Diversity Array) and imputed with curated 1KG as the reference panel where corresponding deep-coverage (>30X) PCR-free whole genome sequencing data were used as a truth call set to calculate sensitivity and specificity. The arrays were also imputed on the Michigan Imputation Server with 1KG as the reference panel.

Within the cohort, four different ancestries were represented - Non-Finnish Europeans (NFE), East Asians (EAS), South Asian (SAS), African (AFR), together with the results for three arrays of undetermined ethnicity (NA).

**Table 1.** Sensitivity and specificity of SNP and INDEL imputation from the Broad Imputation Pipeline (Broad) and Michigan Imputation Pipeline (Michigan) with the curated 1000Genomes reference panel when compared to matched whole genome data for 42 AoU v1 samples.

| Ethnicity | SNP |  |  |  | INDEL |  |  |  |
| --- | --- | --- | --- | --- | --- | --- | --- | --- |
|  | Mean Sensitivity (95% CI) |  | Mean Specificity (95% CI) |  | Mean Sensitivity (95% CI) |  | Mean Specificity (95% CI) |  |
|  | Broad | Michigan | Broad | Michigan | Broad | Michigan | Broad | Michigan |
| NFE<br>(non Finnish European) | 98.5%<br>(98.4 - 98.6) | 98.5%<br>(98.4 - 98.6) | 99.8%<br>(99.8 - 99.9) | 99.8%<br>(99.8 - 99.9) | 97.15%<br>(97.0 - 97.3) | 97.1%<br>(97.0 - 97.3) | 99.4%<br>(99.4 - 99.4) | 99.4%<br>(99.4 - 99.4) |
| EAS<br>(East Asian) | 98.0%<br>(97.1 - 98.8) | 98.0%<br>(97.1 - 98.8) | 99.8%<br>(99.8 - 99.8) | 99.8%<br>(99.8 - 99.8) | 96.4%<br>(95.5 - 97.3) | 96.3%<br>(95.5 - 97.0) | 99.2%<br>(99.1 - 99.4) | 99.3%<br>(99.0 - 99.5) |
| SAS<br>(South Asian) | 97.9%<br>(97.9 - 98.0) | 97.9%<br>(97.9 - 97.9) | 99.8%<br>(99.8 - 99.8) | 99.8%<br>(99.8 - 99.8) | 96.4%<br>(96.3 - 96.5) | 96.3%<br>(96.2 - 96.3) | 99.2%<br>(99.2 - 99.2) | 99.3%<br>(99.2 - 99.3) |
| AFR<br>(African) | 97.3%<br>(97.3 - 97.4) | 96.8%<br>(96.7 - 96.8) | 99.6%<br>(99.5 - 99.7) | 99.5%<br>(99.4 - 99.5) | 95.7%<br>(95.7 - 95.8) | 95.3%<br>(95.2 - 95.3) | 98.7%<br>(98.6 - 98.9) | 98.7%<br>(98.6 - 98.9) |

Broad imputation pipeline sensitivity for SNPs is >97% and INDELs >95% for all ethnicities. Similarly, specificity for SNPs from the Broad imputation pipeline are above 99% and the specificity for INDELS is >98%. Results were highly concordant with those returned by the remote server at Michigan.

##### Performance evaluation of different input material types

To assess the performance of specimens derived from both saliva and whole blood a set of 20 matched blood saliva pairs were run through the GDA genotyping process and the resulting VCFs were imputed using the Broad pipeline to be compared against results for matched blood derived whole genome data. The Pearson correlation between sensitivity and specificity of blood and saliva derived samples are equal to 100% and 100%, respectively. For the same pairs, the Welch two-sample t-test statistic is 0.997 and 0.987, respectively. There is no significant difference between the different input sample types.

##### Imputation repeatability and reproducibility

Imputation pipeline repeatability was assessed by repeating imputation of a cohort of 1000 GSA arrays ten times over the course of two weeks and was found to be 100% concordant. Imputation pipeline precision (reproducibility) was also tested on technical replicates. Three individual samples derived from saliva were each genotyped six times, followed by an imputation in a cohort of all saliva derived samples. In each set of technical replicates all pairs and variants in each pair were compared (making a total of 45 pairs for which genotypes were compared). Reproducibility is measured using Jaccard scores. “Reproducibility over variants” was calculated only over sites where at least one of the two replicates in a pair calls a non hom-ref genotype and was found to be 99.91% (95CI: 99.89-99.93) for SNPs and 99.87% (95CI: 99.85-99.90) for InDels. “Reproducibility over all sites” was calculated over all genotyped sites, including sites genotyped as hom-ref in both replicates and was found to be 100% (95CI: 100-100) for both SNPs and InDels.

##### Imputation performance as a function of variant frequency

Because we expect accuracy to be impacted by the frequency of a variant in the population (rare variants are less likely to be in the reference panel and therefore less accurately imputed) we further subdivided the performance assessment by allele frequencies on two cohorts: 42 AoU v1 arrays and 20 blood-saliva pairs of GDA arrays. Accuracy of imputation of variants as a function of population allele frequency performed as expected with rare variants in the population not being as accurately represented. Imputation is more accurate for variants that are more frequently observed in the population (≥0.1 allele frequency). This is predicted to have a low impact on the accuracy of PRS calculations from imputed variants as PRS scores are also typically derived from common variants.

##### Impact of genotyping array call rate on imputation performance

The impact of call rate on the imputation was assessed by generating a downsampled series of 42 arrays, each with call rates of 90%, 95%, 97% and 98%. Pearson correlation values for SNPs and INDELs were calculated across bins of allele frequencies, assessed against gnomAD common variants (AF >0.1), for the cohorts with downsampled call rates. Call rates below 95% were found to produce suboptimal results. At this rate the mean R^2^ dosage score for sites with AF ≥0.1 was found to be 0.98% (95CI: 0.98-0.98) for both SNPs and InDels compared to 0.99% for call rates of 97% and 98%.

##### Impact of imputation batch size on performance

Batch size effect of the imputation pipeline was assessed by imputing and analyzing arrays in a cohort of size 1000 (randomly chosen), ten cohorts of size 100 (non-overlapping subsets of the 1000 cohort), and ten cohorts of size 10 (non-overlapping subsets of one of the 100 cohorts). Pearson correlations of dosage scores were calculated across bins for allele frequencies (assessed against gnomAD) for smaller cohorts versus larger cohorts. The data show that imputation is highly correlated across batch sizes with batches down to as few as 10 samples producing acceptable performance. The mean R^2^ correlation of dosage scores for sites with allele frequency greater or equal to 0.1 is above 0.97 in all cases both for SNPs and INDELs and increases to 0.98 for the larger studied cohorts. Increasing batch sizes produces very slight improvements in imputation but these are not significant and the choice of imputation batch size (above or equal to 10 samples) can be made on practical and operational grounds

#### Broad PRS Pipeline Overview

The polygenic risk score (PRS) pipeline begins by calculating a raw score using plink2 (https://www.cog-genomics.org/plink/2.0/). For each condition, effect alleles and weights are defined for a set of genomic sites stored in a weights file. At each site, the effect allele dosage observed in the imputed vcf is multiplied by the effect weight in the weights file. The raw score is the sum of these products over all the specified sites.

##### Validation of technical and analytical performance of the PRS pipeline

For each of the 10 conditions chosen by the consortium for clinical return, a validation study was performed to assess the technical and analytical performance as well as to verify the association between score and disease risk.

##### PRS Pipeline Accuracy

Accuracy of the pipeline was determined by calculating the Pearson correlation between PRS scores calculated from 70 specimens imputed from GDA array data and PRS scores of corresponding deep-coverage PCR-free whole genome sequencing data (used as a truth call set).

##### Input Material Performance

Accuracy of PRS scoring when different sample types (blood or saliva) are used as inputs was determined by comparing the PRS scores from matched blood and saliva pairs collected from 20 individuals.

##### PRS pipeline repeatability

PRS pipeline repeatability was assessed by running the pipeline on the same dataset of 70 imputed GDA arrays ten times over the course of two weeks (without call caching). Scores generated from the different processing runs were compared to determine if there are any differences observed for a given PRS when the pipeline is run at different times.

##### PRS pipeline reproducibility

PRS pipeline precision (reproducibility) was assessed using three samples each run 6 times end-to-end and then compared in a pairwise manner. The z-score standard deviation is used as a measure of variability.

##### PRS site representation

The SNP weight sites that are not called during genotyping or imputation were determined. These are sites not present in the intersection of an imputed GDA array and the reference panel. Ideally, all sites required for PRS calculation are present either as genotyped or imputed sites; however, in practice a small number of sites are not present due to differences in the data used to create the score and the specific array and imputation reference panel used in this study.

**Table 2.** Validation measures summary.

|  | PRS Accuracy: Pearson Correlation between PRS from array and WGS* | PRS Pipeline Performance concordance between blood and saliva input types. | PRS Precision: PRS pipeline repeatability | PRS Precision: PRS pipeline reproducibility | PRS Limit of Detection: Score site missingness |
| --- | --- | --- | --- | --- | --- |
| Asthma | 99.3% | 100.0% | 100% | 0.002 | 0.69% |
| Atrial Fibrillation | 98.6% | 100.0% | 100% | 0.001 | 1.20% |
| Breast Cancer | 93.0% | 100.0% | 100% | 0.004 | 0.32% |
| Chronic Kidney Disease | 98.3% | 100.0% | 100% | 0.0001 | 0.69% |
| Coronary Heart Disease | 98.2% | 100.0% | 100% | 0.001 | 0.46% |
| Hypercholesterolemia | 95.9% | 100.0% | 100% | 0.005 | 1.20% |
| Obesity/BMI | 99.5% | 100.0% | 100% | 0.002 | 0.70% |
| Prostate Cancer | 96.4% | 100.0% | 100% | 0.0006 | 2.97% |
| Type 1 Diabetes | 99.5% | 100.0% | 100% | 0.0001 | 2.97% |
| Type 2 Diabetes | 98.8% | 100.0% | 100% | 0.001 | 0.70% |

PRS pipeline accuracy is assessed as the pearson correlation between scores derived from PCR-free WGS and those derived from imputed genotyping data (GDA) in 70 specimens. Pearson correlation shown is the mean correlation across all ancestry groups tested. PRS pipeline performance as a function of sample input was assessed by comparing the scores from 20 matched blood and saliva pairs. PRS pipeline precision (repeatability) is the measure of concordance in PRS scores calculated using the same data from 70 specimens, run through the pipeline 10 times over the course of two weeks. PRS pipeline precision (reproducibility) was assessed using three samples each run 6 times end-to-end and then compared in a pairwise manner. The z-score standard deviation is used as a measure of variability. PRS site representation is a measure of the percentage of the original score sites are missing from the final imputed dataset.

##### Performance verification using eMERGE I-III cohort

A cohort of samples with known phenotypic information was used to verify the relationship between polygenic risk score as determined by our pipeline and disease risk. For conditions where cases and controls could be identified in the eMERGE I-III cohort we determined performance using metrics outlined in the ClinGen working group recommendations (Wand et al.). Specifically, we determined the PRS distributions for cases and controls, we examined the impact of ancestry adjustment on the distributions (Fig), and we examined the relationship between observed and predicted risk. There are some limitations to this analysis: i) The eMERGE I-III dataset being used for this analysis was generated from different array platforms and was imputed with a different pipeline including a different version of 1KG reference panel than the one currently implemented; ii) The eMERGE I-II-III imputed dataset does not include variants from Chromosomes X or Y. For these reasons, the PRS disease association analysis represents a verification of the clinical validation performed by eMERGE IV condition leads rather than the quantitative measure of the impact of the score on risk. The clinical associations (odd ratios) that are reported on the clinical report for each condition were independently determined by eMERGE IV disease-specific expert teams.

##### Validation of pipeline and ancestry adjustment in original case control cohorts

The final pipeline was made available to computational scientists at each of the eMERGE IV disease-specific expert teams who had access to appropriate case control cohorts. These groups confirmed the performance of the final pipeline on their cohorts. The odds ratios for each condition that are reported on the clinical reports come from these cohorts rather than the eMERGE cohort for the reasons described above.

### B. PRS Ancestry Calibration Overview

#### PCA method description

For a polygenic risk score which is a sum of SNP effects (linear weights), the central limit theorem states that the distribution of scores in a homogenous population will tend towards a normal distribution as the number of SNPs becomes large. When two different homogenous populations are randomly mixed, the additive property of prs leads the resulting distribution to be similarly normally distributed, with mean and variance depending on the mean and variance of the original homogenous populations. We can therefore model the distribution of prs scores as being normally distributed, with mean and variance being functions of genetic ancestry.

Practically, we implement this as

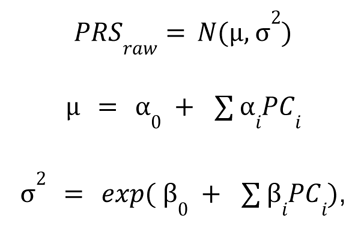

with genetic ancestry being represented by projection into principal component space. The **α** and **β** parameters are found by jointly fitting them to a cohort of training data. This fit is performed by minimizing the negative log likelihood:

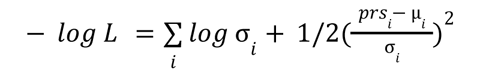

where ι runs over the individuals in the training cohort, *prs_j_* is the i’th individuals raw prs score, and µ and σ are calculated using Eq X by projecting the i’th individual into PC space. Note that, due to the simplicity of the model, overfitting is unlikely to be a problem, and so no regularization or other overfitting avoidance technique is implemented. An individual’s PRS z-score can then be calculated as

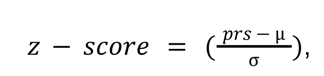

where µ and σ have again been calculated based on the specific individuals projection into PC space. In this way, once the model has been trained, the z-score calculation is fully defined by the fitted model parameters, and z-scores can be calculated without needing additional access to the original training cohort.

#### Generating trained models from All of Us data

Generating the trained models consisted of three steps: 1. Selecting the training cohort. 2. Imputation of the training cohort. 3. Training the models on the training cohort. A test cohort was also generated in order to test the performance of the training.

Ancestry balanced training and test cohorts were generated by subsampling from an initial cohort of around 100,000 All of Us samples. For the purposes of balancing the cohort, each sample was assigned to one of the five 1KG Super Populations. Principal component analysis was first performed on a random subset of 20,000 samples. 1KG samples were projected onto these principal components, and a support vector machine (SVM) was trained on 1KG to predict ancestry. The SVM was then used to assign 108,000 AoU samples to one of the five 1KG Super Populations. A balanced training cohort was selected based on these predicted ancestries, and principal components were recalculated using this balanced training cohort. A similarly balanced test cohort was selected based on ancestries estimated from projection on the training set PCs. The resulting breakdown of the cohorts by estimated ancestry is shown in Table 3.

**Table 3.**
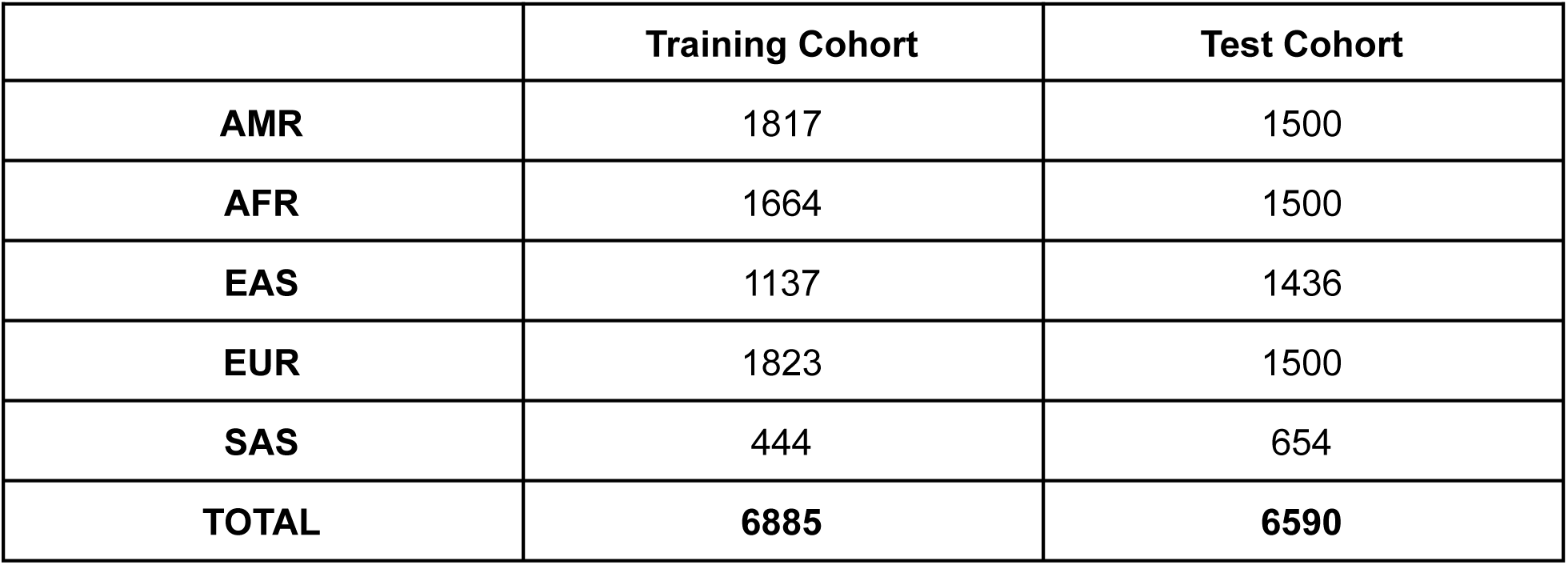

|  | Training Cohort | Test Cohort |
| --- | --- | --- |
| **AMR** | 1817 | 1500 |
| **AFR** | 1664 | 1500 |
| **EAS** | 1137 | 1436 |
| **EUR** | 1823 | 1500 |
| **SAS** | 444 | 654 |
| **TOTAL** | **6885** | **6590** |

Both the training and testing cohorts include a number of individuals with highly admixed ancestry. Admixture was quantified using the tool Admixture (Alexander *et al. PMID: 19648217)* with 5 ancestral populations. The resulting admixtures of each cohort are shown in Figure 1, and the most common admixed ancestries in each cohort are summarized in Table 4.

Each cohort was imputed using the imputation pipeline described above, with 1KG as the reference panel. By keeping the imputation pipeline identical to the pipeline used for the eMERGE dataset, and because the AoU dataset uses the same GDA array as the eMERGE dataset, any potential biases resulting from differing data production and processing methods were removed. The training cohort was scored for each of the ten conditions, and model parameters were fit by minimizing the negative log likelihood as described. The test cohort was then used to evaluate the generalizability of these model parameters.

**Figure 1.**
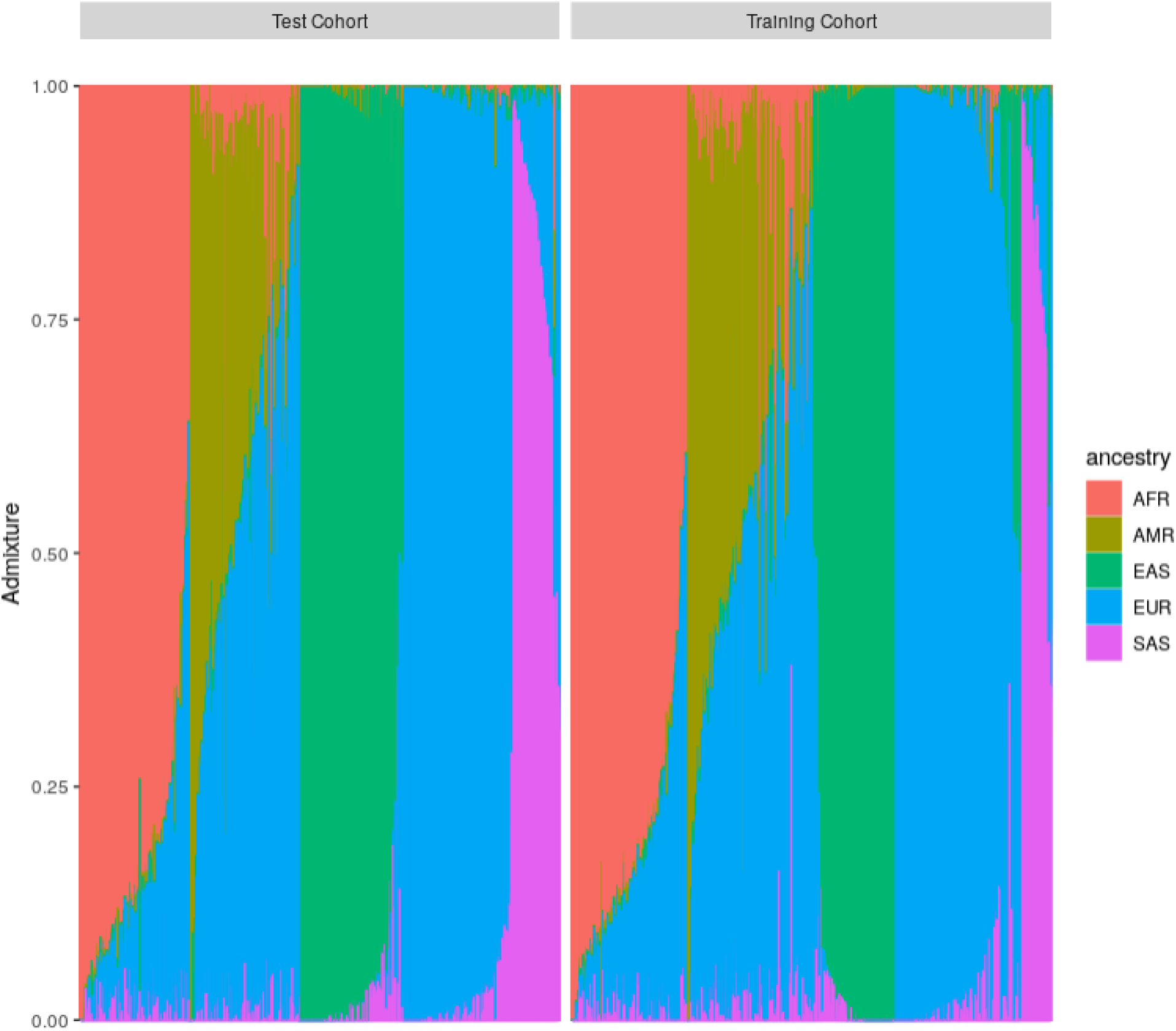

**Table 4.** Admixed ancestries are defined as ancestries for which an individual’s admixture fraction is greater than 20%. For example, an individual who is indicated by admixture to be

| Admixed Ancestry | Training Cohort | Test Cohort |
| --- | --- | --- |
| **AFR-EUR** | 590 | 556 |
| **AMR-EUR** | 1238 | 883 |
| **EAS-EUR** | 236 | 102 |
| **EUR-SAS** | 191 | 229 |

45% AFR, 37% EUR, 12 % AMR, 5% EAS, 1% SAS would be included in the AFR-EUR row of this table.

#### Performance on Test Cohort

Figure 2 the distribution of calibrated z-scores in the test cohort using the parameters fit in the training cohort. As can be seen, all ancestries show the intended standard normal distribution of calibrated scores.

**Figure 2.**
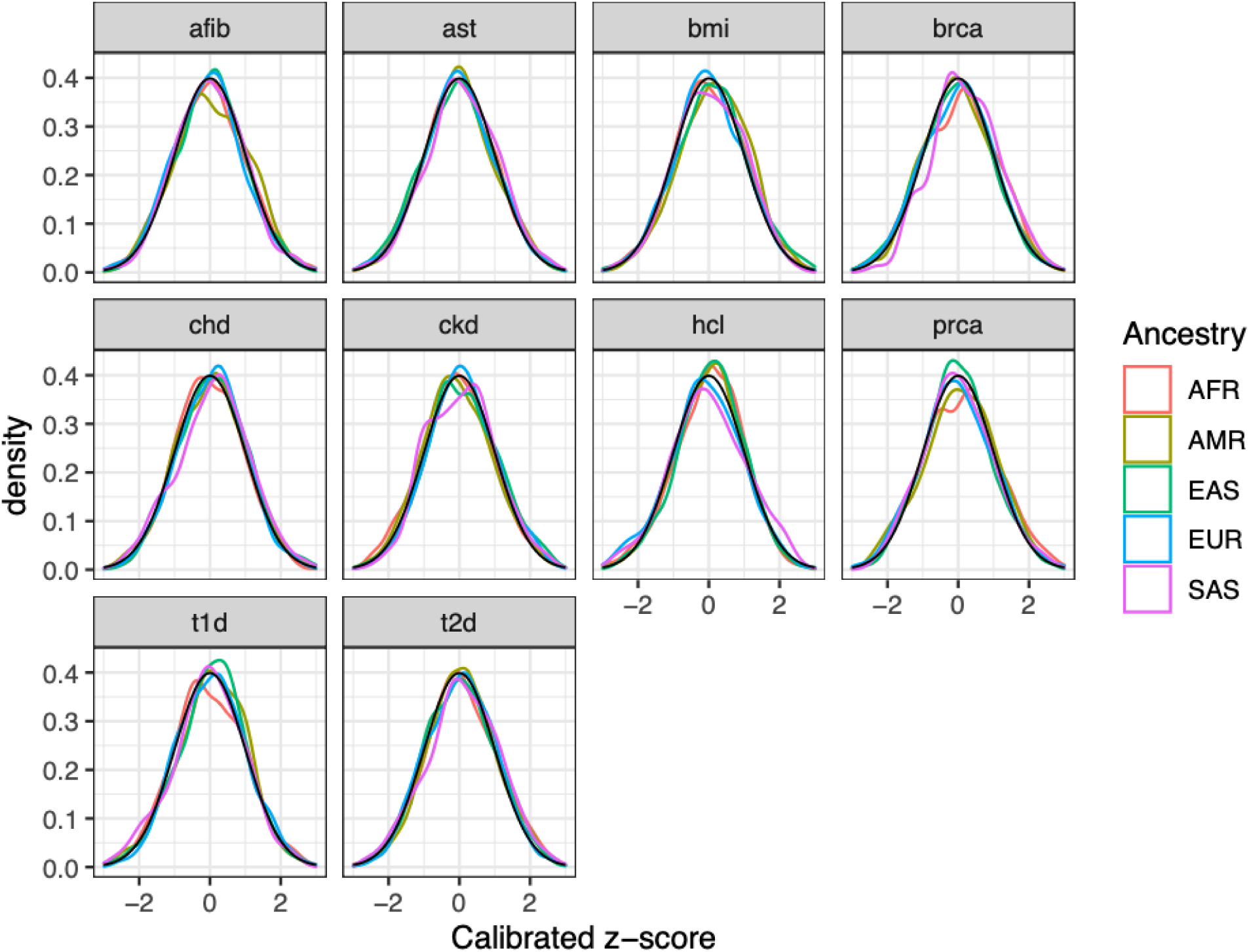

One of the main improvements of this method over previous methods is the inclusion of an ancestry dependent variance in addition to the ancestry dependent mean. The importance of this is shown for the Hypercholesterolemia PRS in Figure X below. As can be seen, the variance of this score differs significantly across ancestries, so that a method which only fits the mean of the distribution as ancestry dependent can result in z-score distributions which have been attenuated towards zero or expanded away from zero for some ancesties. By also treating variance as ancestry dependent, this method results in z-score distributions which are more standardized across ancestries.

**Figure 3.**
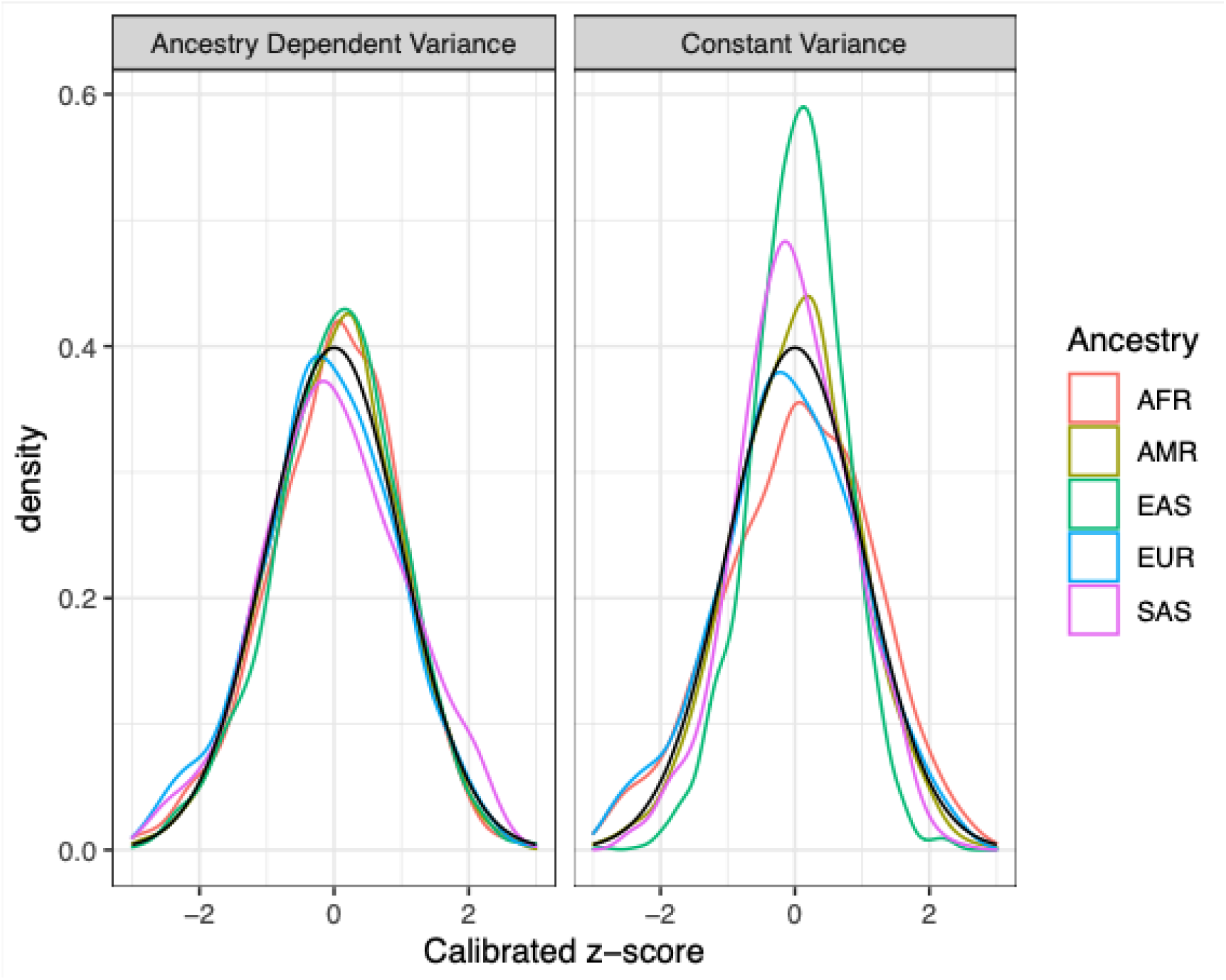
Hypercholesterolemia PRS calibrated z-scores of training cohort. Note the improvement when an ancestry dependent variance is used over a constant variance method.

In addition to improving calibration across ancestries, this method can improve calibration within ancestries, particularly for highly admixed individuals. An example of this can be seen in Figure 4. As can be seen, because no ancestry group is homogenous, when individuals are compared directly to other individuals in their assigned population group, a dependence between admixture fraction and PRS score can result. This dependence is removed by the described PCA calibration method, and the resulting calibrated PRS scores are independent of admixture fraction.

**Figure 4.**
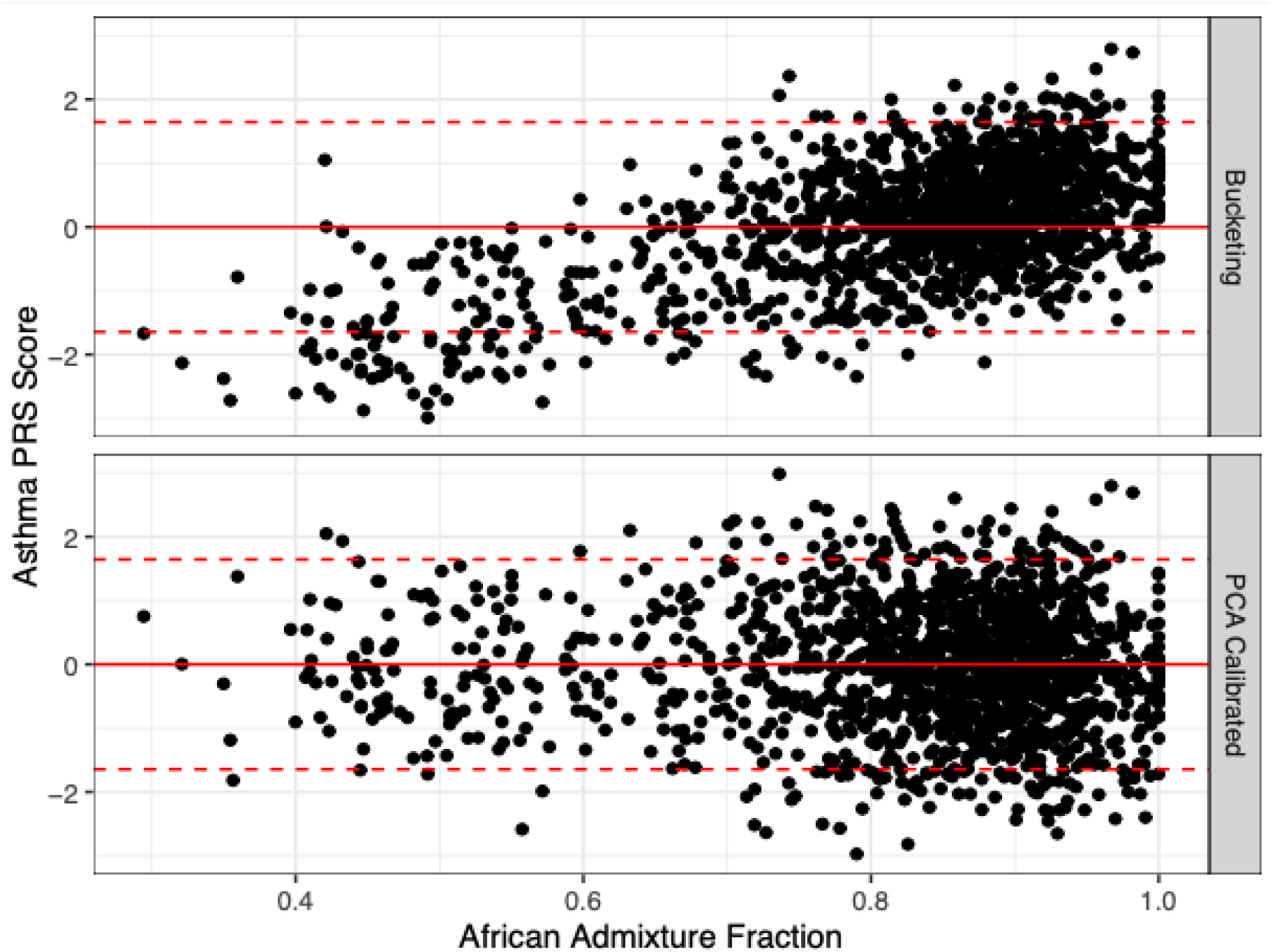
PRS z-score as a function of African Admixture Fraction, for individuals of African ancestry. In the “Bucketing” method, a z-score is calculated by comparing to the mean and variance of all individuals of African ancestry in the cohort. The “PCA Calibrated” method is the method described above. Note the dependence on admixture fraction in the “Bucketing” method, which has been removed in the “PCA Calibrated” method

**Supplemental Table 1.**
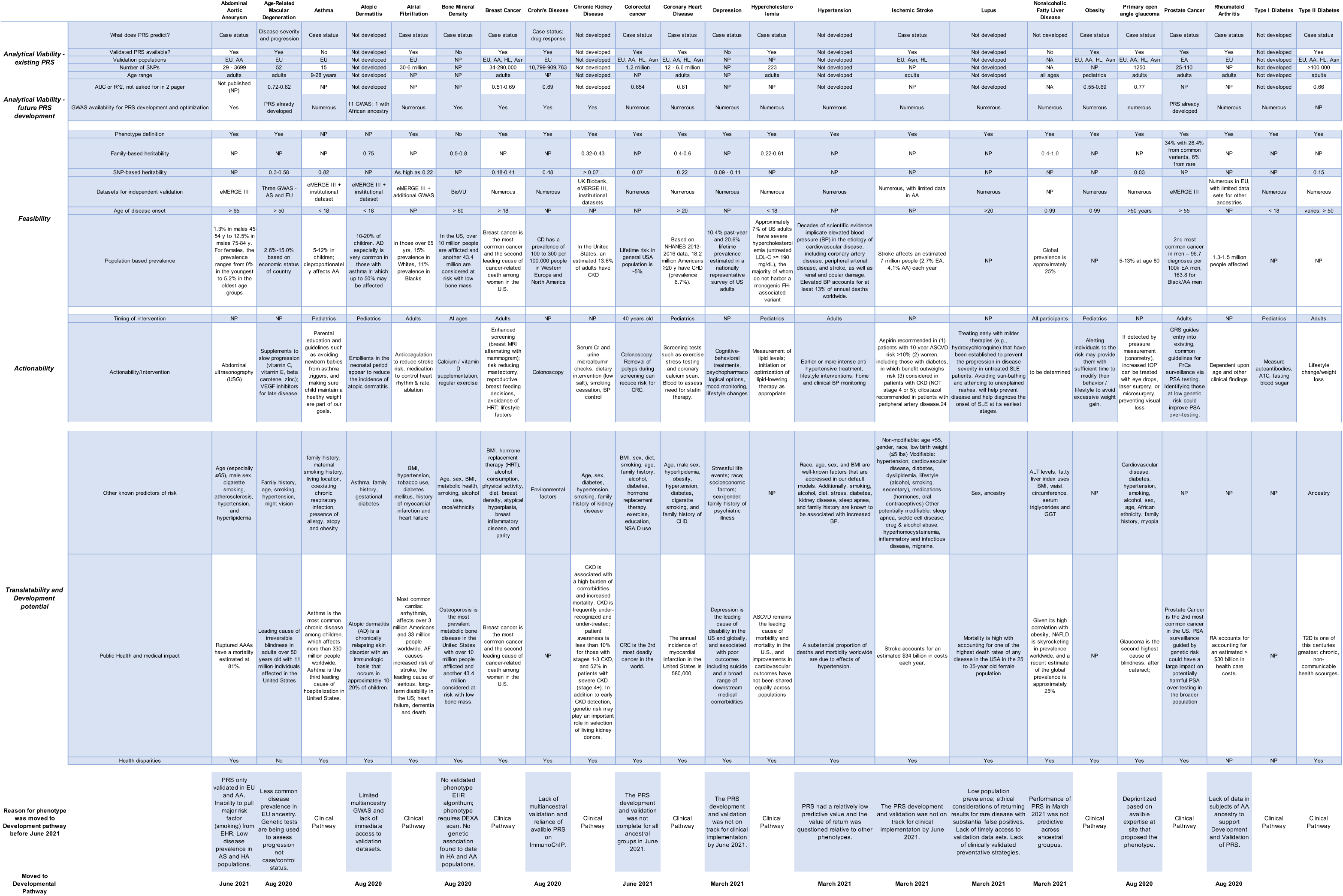

**Supplemental Table 2.**
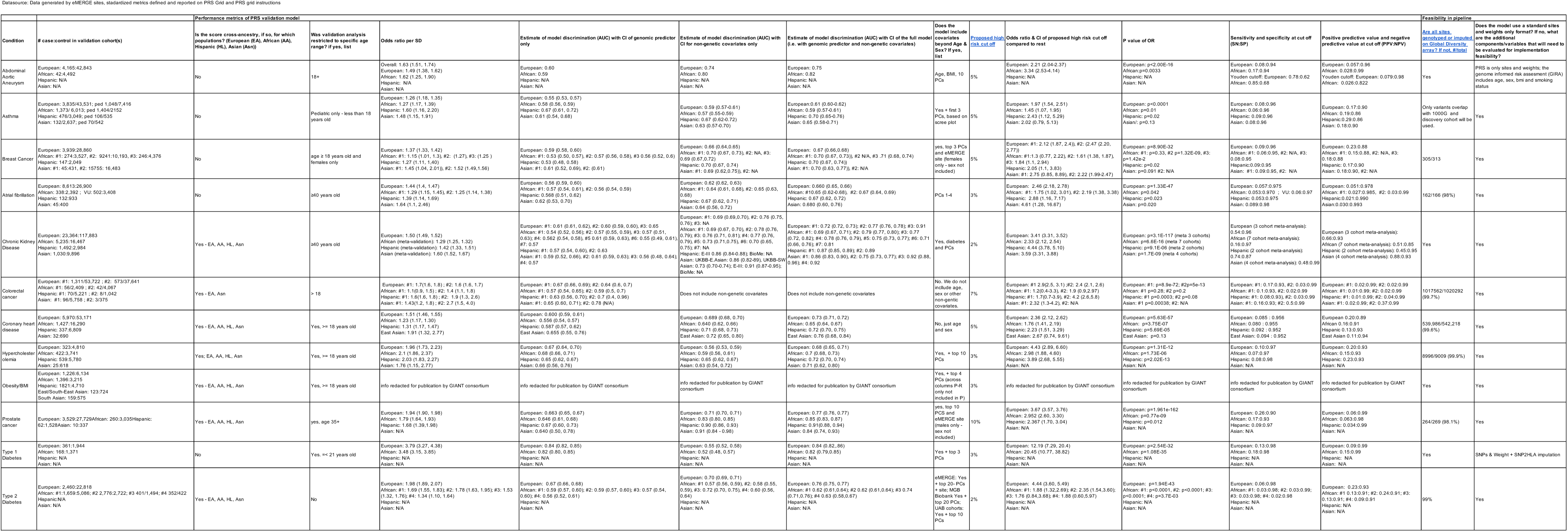

**Supplemental Table 3.**
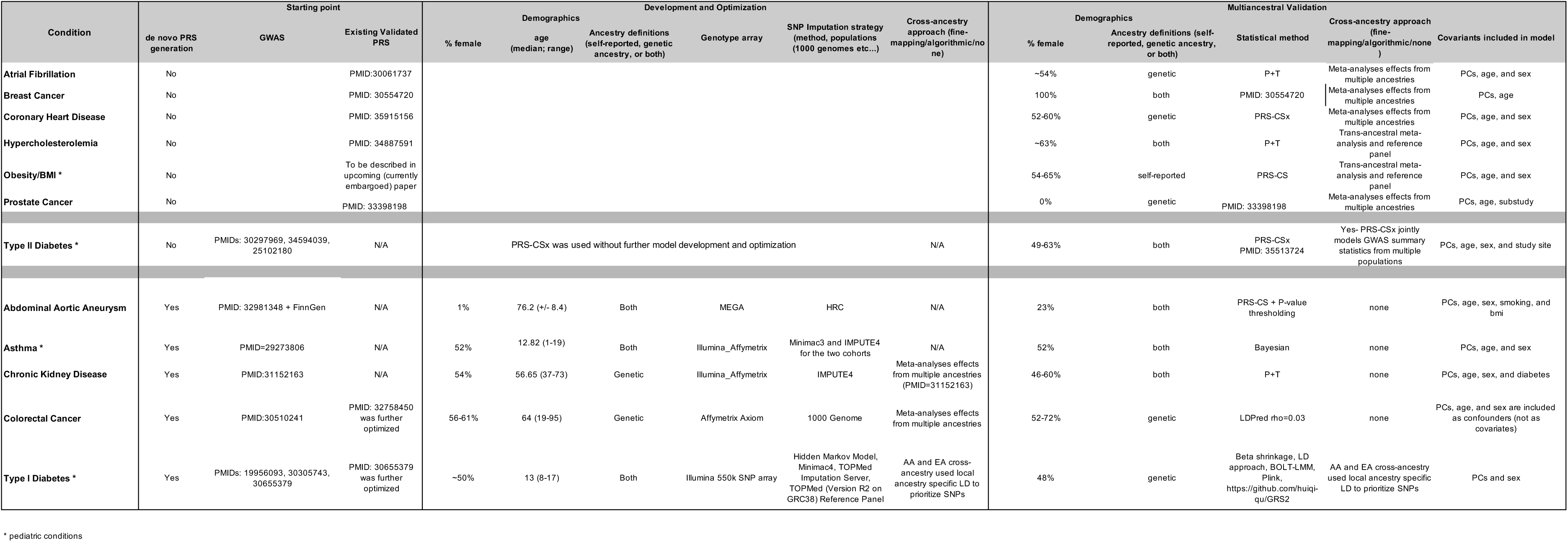

